# Geographic variation in potentially preventable hospitalisations in Indonesia: a multilevel analysis using National Health Insurance Sample Data

**DOI:** 10.1101/2025.11.12.25340113

**Authors:** Dewi Amila Solikha, Danielle C. Butler, Rosemary J. Korda, Matthew Kelly

**Affiliations:** National Centre for Epidemiology and Population Health, Australian National University, Canberra, ACT, Australia; Directorate for Public Health and Nutrition, Ministry of National Development Planning, Jakarta Pusat, DKI Jakarta, Indonesia

**Keywords:** health equity, health services, quality in health care, hospitalisation, Indonesia, primary health care

## Abstract

**Objectives:** Indonesian data on potentially preventable hospitalisations (PPH), a widely used indicator for measuring primary health care (PHC) quality, are lacking. This study aimed to describe Indonesia’s PPH rates and to quantify variation in PPH across districts/cities and their associations with individual sociodemographic characteristics.

**Design and setting:** Cross-sectional study conducted in Indonesia using the 2023 National Health Insurance Sample Data.

**Participants:** Sample of 2.3 million individuals, representing 266.8 million national health insurance−*Jaminan Kesehatan Nasional* (*JKN*)−enrolees.

**Outcome measures:** PPH overall and by type: vaccine-preventable, acute, and chronic.

**Methods:** We quantified crude and age-standardised rates per 10,000 *JKN* enrolees. We applied multilevel negative binomial regression and estimated: 1) median rate ratios (MRRs) to quantify variation in PPH across districts/cities, unadjusted and adjusted for sociodemographic characteristics, and 2) incidence rate ratios (IRRs) to quantify associations between PPH and sociodemographic characteristics.

**Results:** There were 4.83 million overall PPHs in 2023, an overall rate of 180.9 per 10,000 *JKN* enrolees (acute: 94.8, chronic: 93.6, vaccine-preventable: 2.8). Overall PPH rates were generally highest in districts/cities in Java-Bali, Sulawesi, Sumatera, Kalimantan, lower in Nusa Tenggara, Maluku, and lowest in Papua. Variation in PPH across districts/cities was similar for all PPH types (MRRs overall: 1.62, vaccine-preventable: 1.83, acute: 1.72, chronic: 1.53) and persisted after adjusting for sociodemographic characteristics (MRRs of 1.52, 1.82, 1.61, 1.41, respectively). Higher PPH was strongly associated with older age (55−64 and 65−74 years, particularly chronic) and young age (0−14 years, vaccine-preventable, acute), modestly associated with being married or divorced (acute, chronic, overall), and being non-subsidised (all types), and weakly associated with sex (all types).

**Conclusions:** PPHs varied across districts/cities as well as by sociodemographic characteristics. While PPH has potential for measuring PHC quality in Indonesia, context-specific interpretation is required as lower PPH may reflect limited accessibility to hospital-care rather than better PHC quality.

**Strengths and limitations of this study:**

⇒ This is the first study to describe potentially preventable hospitalisations (PPH) in Indonesia, one of the most populous countries in the Asia-Pacific region and globally, using a large-scale sample from routine national health insurance (*Jaminan Kesehatan Nasional or JKN,* one of the world’s largest health insurance schemes) data.
⇒ We applied two-level multilevel analysis to account for the hierarchical structure of the data (individuals nested within districts/cities).
⇒ Our study did not capture all PPH in Indonesia as around 28% of hospitalisations are not through *JKN* scheme.
⇒ Indonesia has not yet developed its own list of conditions for defining PPH. We modified lists of conditions from other countries, which may not fully reflect the Indonesian context.

## INTRODUCTION

Indonesia has undergone major primary health care (PHC) policy reforms over recent decades. These reforms include enhancing the capacity and capability of PHC services as well as strengthening preventive and promotive activities [1,2]. The Medium-Term Development Plan (*RPJMN*) 2025−2029 further reflects the Indonesian Government’s commitment to continue investing in PHC to ensure equitable access to high-quality care across geographic locations over the next five years [3].

Indonesia’s PHC system comprises public and private providers operating in more than 23,000 PHC facilities across the country [4]. These facilities deliver a range of outpatient services, basic short-stay inpatient services, and public health functions such as health promotion and disease prevention [5]. PHC serves as the first point of contact, particularly for low-income populations in remote areas [6]. Under Indonesia’s health insurance program, *Jaminan Kesehatan Nasional* (*JKN*), PHC plays a gatekeeping role within a tiered referral system, whereby access to secondary hospital care requires prior assessment and referral from PHC providers [5].

Indonesia operates a decentralised health system across 38 provinces and 514 districts/cities, with responsibility for managing healthcare resources including infrastructures, financing, workforces, and medicines, being delegated to subnational (provincial and district/city) governments. As PHC services are delivered at the subdistrict level, district/city policies directly affect PHC operations [7–9]. Consequently, variations in capacity and policy priorities across districts/cities influence PHC performance nationally. Monitoring and evaluating ongoing PHC policy reforms and investments are essential to assess progress toward intended objectives, particularly achieving equitable access to high-quality PHC services, and to inform appropriate policy decisions.

The WHO/UNICEF PHC measurement framework [10] outlines a range of indicators for measuring PHC performance, including quality of care. One such indicator is potentially preventable hospitalisations (PPH), for which the most common definition is hospitalisations that could potentially have been prevented through timely and effective PHC [11]. International literature shows that PPH is widely used as a practical measure of PHC quality [10,12–17], although the extent to which PPH accurately reflects the quality of PHC may vary depending on the healthcare system and country context [18–21].

A recent scoping review on PHC performance measurement in Indonesia found that although indicators for quality of care have been explored, no studies to date have specifically examined PPH [22] or assessed the extent of its geographic variation. Therefore, this study aimed to 1) describe PPH rates (overall, and by PPH type, sociodemographic characteristics, disease category, and districts/cities) among *JKN* enrolees in Indonesia in 2023, and 2) quantify variation in PPH across districts/cities and associations with individual sociodemographic characteristics.

## METHODS

### Study population and setting

We conducted a cross-sectional study using data from the most recent (2023) National Health Insurance Sample Data (hereafter referred to as NHISD), a large-scale routinely collected dataset provided by *Badan Penyelenggara Jaminan Sosial (BPJS) Kesehatan*, Indonesia’s Social Security Administering Body for Health. NHISD contains anonymised individual-level healthcare utilisation records under the *JKN* scheme and is publicly accessible upon request to *BPJS Kesehatan*.

NHISD includes a 1% stratified random sample of *JKN* enrolees. The primary sampling unit is PHC facilities. Families registered at each PHC facility were stratified into three groups: 1) never utilised any healthcare services, 2) utilised PHC services, and 3) utilised both PHC and hospital services. Ten families from each stratum at each PHC facility were randomly selected, with all family members included. Oversampling was applied to those who used health services, resulting in unequal selection probabilities. To account for this potential bias, we applied weights provided in the dataset.

The study population, drawn from NHISD, included *JKN* enrolees who were 1) active: those who regularly paid premium and had full access to health services, or 2) passive: those who had missed premium payments and may have had limited access to health services but were assumed to have possibly used services during 2023 once they became active again, as NHISD did not provide information on the duration of passive status. We excluded: 1) individuals with duplicate *JKN* memberships, 2) those residing outside Indonesia, 3) those who died before 2023, and 4) residents of *Deiyai* District (Papua), where no hospitals were contracted under *JKN* in 2023, resulting in no hospital utilisations records in NHISD 2023. We did not exclude those who died in 2023, under the assumption that they may have accessed health services during that year prior to death (exact dates of death were not available in NHISD).

### Data

Sociodemographic and health service utilisation data were obtained from the 2023 NHISD. We used three of the five relational sub-datasets: 1) the membership sub-dataset, which contains sociodemographic information including age, marital status, sex, type of *JKN* membership, and area of residence; 2) the hospital sub-dataset, which records all hospital inpatient visits along with primary diagnoses; and 3) the secondary diagnosis sub-dataset, which documents additional diagnoses recorded during the same hospital admissions. All three sub-datasets were linked using individual identification numbers.

### Variables

#### Outcome of interest

Our outcome was PPH. To define PPH, we used International Classification of Diseases, 10^th^ revision (ICD-10) codes to identify conditions classified as PPH, based primarily on the primary diagnosis. The initial list for defining relevant conditions was modified from a study of PPH in Vietnam which used health insurance claims data [23], considered contextually relevant to Indonesia given both countries’ epidemiological profiles of rising non-communicable diseases alongside persistent communicable diseases burdens [24]. We further refined the list using Brazil’s list [25] including tropical diseases, particularly malaria, that are prevalent in Indonesia [26] and Australia’s list [17] which provided detailed inclusion and exclusion criteria. The full list of conditions used to define PPH for this study is provided in **Supplemental table 1**. For each individual, we summed overall PPH events in 2023. We also summed PPH events according to type, based on international practice [15–17]: vaccine-preventable, acute, and chronic PPH. In calculating overall PPH, each admission was counted only once even if multiple PPH conditions (vaccine-preventable, acute, and chronic) were reason for admissions, consistent with Australia’s approach [27].

#### Sociodemographic characteristics

Individual sociodemographic characteristics derived from NHISD included age (categorised into 0−14, 15−24, 25−34, 35−44, 45−54, 55−64, 65−74, 75+ years), marital status (single/married/widowed), sex (male/female), and type of *JKN* membership (subsidised/non-subsidised group). Individuals in the subsidised group were regarded as poor or near poor, with premiums paid by the government. The non-subsidised group included those who paid their own premiums, encompassing paid and unpaid workers as well as non-workers.

#### Geographic unit of analysis

We used district/city as the unit of area-level context. Districts/cities were assigned to *JKN* enrolees based on their area of residence recorded in NHISD membership sub-dataset. While NHISD includes 514 districts/cities, our analysis covered 513, excluding *Deiyai* District in Papua due to the absence of *JKN*-contracted hospitals in 2023. For interpretability, we grouped the 513 districts/cities into seven regions: 1) Sumatra, 2) Java-Bali, 3) Kalimantan, 4) Sulawesi, 5) Nusa Tenggara, 6) Maluku, and 7) Papua [28].

#### Statistical analysis

We quantified crude PPH rates as the number of PPH events in 2023 divided by the total number of *JKN* enrolees in NHISD for the same year. Rates were calculated overall and separately by type (vaccine-preventable, acute, and chronic PPH), sociodemographic characteristics, and disease category. We then analysed variation in PPH across districts/cities in two steps. We first calculated age-standardised PPH rates by district/city (grouped by region) using direct standardisation with national *JKN* enrolees as the standard population and plotted the results. Both crude and age-standardised PPH rates were calculated using sampling weights from NHISD to account for the sampling design and represent all *JKN* enrolees. Second, we performed two-level multilevel negative binomial models to account for the hierarchical structure of the count data (individuals nested within districts/cities) [29] and to address overdispersion [30]. We applied two model specifications: model 1 included only a random intercept to quantify district/city variation for each outcome (overall PPH and by type) and model 2 included age, marital status, sex, and type of *JKN* membership to adjust for individual sociodemographic characteristics in the variation and to estimate their associations with outcomes.

Multilevel analysis results were presented as the median rate ratio (MRR) and incidence rate ratio (IRR). The MRR was used to estimate variation in PPH across districts/cities. The MRR, calculated based on the variance term (σ^2^), quantifies the median relative change in outcomes when individuals move from one area to another higher risk area, using the formula MRR = exp 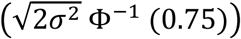 where Φ^−1^ refers to the 75^th^ percentile of a standard normal distribution [29]. We also calculated IRRs and 95% confidence intervals to estimate associations between sociodemographic characteristics and PPH. Missing data for marital status and type of *JKN* membership were addressed using logistic regression imputation. Analyses were conducted in Stata version 19.5.

## RESULTS

### Sample characteristics

After excluding individuals with duplicate data (n=68,125), those residing abroad (n=57,269), those who died before 2023 (n=54,642), and those residing in *Deiyai* District (n=1,609), the final study sample comprised 2,319,606 unweighted or 266,855,314 weighted individuals (**Figure 1**). The study samples’ mean of age was 36.4 years (SD 20.3), 55.3% were married, 51.1% were male, and 62.0% were in the subsidised group (**Table 1**).

**Figure 1.**
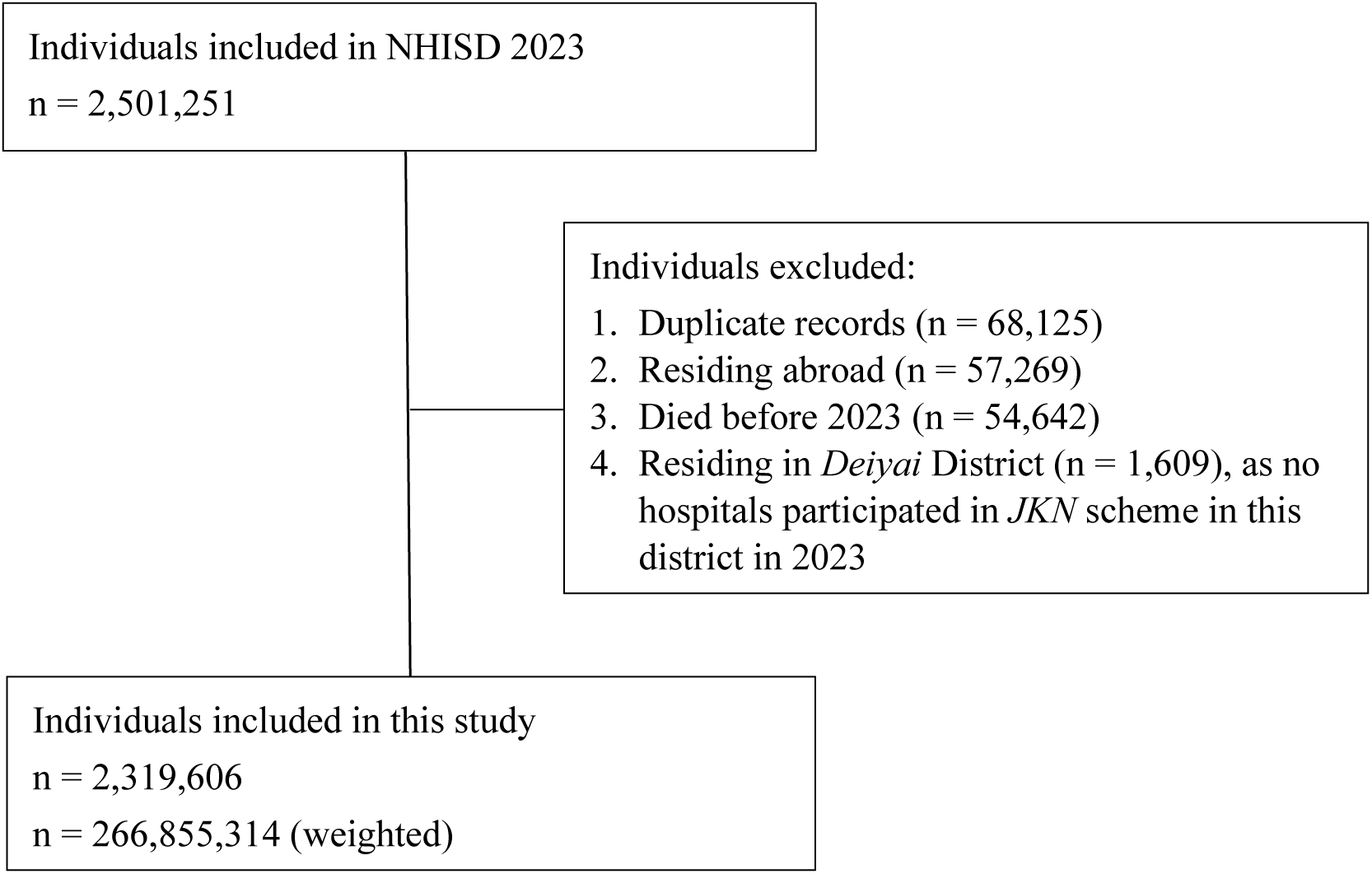
Flow diagram of study population included in this study Each box presents unweighted number of individuals, but the final box shows both unweighted and weighted number of individuals (using sampling weights provided in NHISD 2023). NHISD, National Health Insurance Sample Data; *JKN, Jaminan Kesehatan Nasional* (National Health Insurance); n, number of individuals.

**Table 1.** Sample characteristics: individual sociodemographic characteristics and PPH rates.

| Variable | Study sample:<br>persons in<br>million<br>(%) | Overall PPH |  | Vaccine-preventable PPH<br>Rates per<br>10,000<br>*JKN* enrollees | Acute PPH<br>Rates per<br>10,000<br>*JKN* enrollees | Chronic PPH<br>Rates per<br>10,000<br>*JKN* enrollees |
| --- | --- | --- | --- | --- | --- | --- |
|  |  | Number in<br>million<br>(%) | Rates per<br>10,000<br>*JKN* enrollees |  |  |  |
| Total | 266.85<br>(100) | 4.83<br>(100) | 180.9 | 2.8 | 94.8 | 93.6 |
| Age group |  |  |  |  |  |  |
| 0–14 | 39.93<br>(15.0) | 0.79<br>(16.4) | 198.7 | 5.8 | 180.0 | 14.3 |
| 15–24 | 46.31<br>(17.4) | 0.37<br>(7.6) | 79.4 | 1.3 | 61.8 | 17.2 |

| Variable | Study sample:<br>persons in<br>million<br>(%) | Overall PPH |  | Vaccine-preventable PPH | Acute PPH | Chronic PPH |
| --- | --- | --- | --- | --- | --- | --- |
|  |  | Number in<br>million<br>(%) | Rates per<br>10,000<br>*JKN* enrollees | Rates per<br>10,000<br>*JKN* enrollees | Rates per<br>10,000<br>*JKN* enrollees | Rates per<br>10,000<br>*JKN* enrollees |
| 25–34 | 45.77<br>(17.1) | 0.35<br>(7.3) | 77.5 | 1.6 | 53.7 | 23.4 |
| 35–44 | 44.58<br>(16.7) | 0.47<br>(9.8) | 106.1 | 2.2 | 57.5 | 51.3 |
| 45–54 | 36.61<br>(13.7) | 0.78<br>(16.1) | 211.8 | 3.2 | 81.9 | 145.0 |
| 55–64 | 26.08<br>(9.8) | 1.04<br>(21.6) | 399.2 | 4.4 | 133.8 | 296.7 |
| 65–74 | 15.28<br>(5.7) | 0.71<br>(14.8) | 467.4 | 2.2 | 166.7 | 336.1 |
| 75+ | 12.29<br>(4.6) | 0.31<br>(6.4) | 249.8 | 1.1 | 97.6 | 168.5 |
| Marital status |  |  |  |  |  |  |
| Single | 111.31<br>(41.7) | 1.40<br>(29.1) | 126.2 | 3.3 | 99.4 | 25.6 |
| Married | 147.67<br>(55.3) | 3.07<br>(63.6) | 207.8 | 2.4 | 86.8 | 133.8 |
| Divorced | 7.88<br>(3.0) | 0.35<br>(7.3) | 448.8 | 3.1 | 181.2 | 300.1 |
| Sex |  |  |  |  |  |  |
| Male | 136.24<br>(51.1) | 2.36<br>(48.9) | 173.3 | 3.1 | 87.5 | 92.6 |
| Female | 130.62<br>(48.9) | 2.47<br>(51.1) | 188.8 | 2.4 | 102.4 | 94.7 |
| Type of *JKN* membership |  |  |  |  |  |  |
| Subsidised<br>group | 165.39<br>(62.0) | 2.24<br>(46.4) | 135.4 | 2.1 | 67.7 | 73.6 |
| Non-subsidised<br>group | 101.46<br>(38.0) | 2.59<br>(53.6) | 255.0 | 3.8 | 139.1 | 126.2 |

Rates are crude, calculated per 10,000 *JKN* enrollees using sampling weights provided in NHISD 2023. PPHs were grouped into overall, and by type: vaccine-preventable, acute, and chronic. Columns for each study sample and numbers of overall PPH sum to 100% across categories for each variable. Numbers of study participants with missing data were 231,733 persons (9.9%) for marital status and 23 persons (<0.001%) for type of *JKN* membership, both variables were imputed using regression imputation. There were no missing data for age group or sex.
PPH, potentially preventable hospitalisations; *JKN*, *Jaminan Kesehatan Nasional* (Indonesia's national health insurance scheme).

### PPH rates overall and by type, disease category, and sociodemographic characteristics

Among the study sample, 266.8 million weighted individuals (**Table 1**), we observed 4.83 million overall PPH events, corresponding to a rate of 180.9 per 10,000. This included rates of 94.8 for acute, 93.6 for chronic, and 2.8 for vaccine-preventable PPH. For acute conditions (**Table 2**), dehydration and gastroenteritis (37.67), and bacterial pneumonia (27.14) were the leading causes of PPH. For chronic conditions, diabetes and associated complications (20.96), hypertensive complications (19.72), cerebrovascular disease (17.72), and tuberculosis (8.92) were the primary contributors. Highest PPH rates (**Table 1**) were observed in older age 55−64 and 65−74 years (chronic and overall PPH), young age 0−14 years (vaccine-preventable and acute PPH), divorced individuals (acute, chronic, and overall PPH), females (acute, chronic, and overall PPH), and non-subsidised group (all PPH types).

**Table 2.** Distribution of PPH rates per 10,000 *JKN* enrolees for each disease category and sex.

| Disease category of PPH |  | Male | Female | Total |
| --- | --- | --- | --- | --- |
| Vaccine-preventable disease |  |  |  |  |
| 1 | Influenza and pneumonia (vaccine-preventable) | 0.02 | 0.00 | 0.01 |
| 2 | Other vaccine-preventable diseases | 3.13 | 2.39 | 2.77 |
| Acute disease |  |  |  |  |
| 3 | Bacterial pneumonia (not vaccine-preventable) | 27.33 | 26.94 | 27.14 |
| 4 | Cellulitis | 0.00 | 0.00 | 0.00 |
| 5 | Dehydration and gastroenteritis | 33.20 | 42.34 | 37.67 |
| 6 | Dental conditions | 1.32 | 1.09 | 1.21 |
| 7 | Ear, nose, throat (ENT) | 9.17 | 7.84 | 8.52 |
| 8 | Gangrene | 6.38 | 6.96 | 6.66 |
| 9 | Pelvic inflammatory disease | 0.00 | 1.91 | 0.93 |
| 10 | Perforated or bleeding ulcer | 2.60 | 2.40 | 2.50 |
| 11 | Pyelonephritis and diseases of the urinary system | 3.57 | 9.38 | 6.41 |
| 12 | Gallstone ileus | 0.00 | 0.00 | 0.00 |
| 13 | Appendicitis complications | 1.36 | 1.49 | 1.43 |
| 14 | Seizure disorders | 3.15 | 2.70 | 2.93 |
| 15 | Malaria | 0.10 | 0.04 | 0.07 |
| Chronic disease |  |  |  |  |
| 16 | Nutritional deficiencies | 0.06 | 0.07 | 0.06 |
| 17 | Tuberculosis | 11.66 | 6.05 | 8.92 |
| 18 | Syphilis | 0.04 | 0.02 | 0.03 |
| 19 | Angina | 6.05 | 4.59 | 5.33 |
| 20 | Congestive heart failure | 6.75 | 5.01 | 5.90 |
| 21 | Hypertensive complications | 19.40 | 20.04 | 19.72 |
| 22 | Rheumatic heart disease | 0.43 | 0.28 | 0.36 |
| 23 | Asthma | 3.43 | 5.57 | 4.48 |
| 24 | Chronic obstructive pulmonary disease (COPD) | 7.60 | 5.95 | 6.79 |
| 25 | Diabetes and associated complications | 15.94 | 26.21 | 20.96 |
| 26 | Iron-deficiency and other anaemia | 0.10 | 0.27 | 0.18 |
| 27 | Cerebrovascular disease | 18.52 | 16.87 | 17.72 |
| 28 | Atrial fibrillation | 1.35 | 1.92 | 1.63 |
| 29 | Dementia | 0.24 | 0.11 | 0.18 |
| 30 | Depressive disorders | 0.13 | 0.22 | 0.17 |
| 31 | Colorectal cancer | 0.88 | 0.67 | 0.78 |
| 32 | Cervical cancer | 0.00 | 0.81 | 0.40 |

Rates are crude, calculated per 10,000 *JKN* enrollees using sampling weights provided in NHISD 2023. PPH by type were grouped into vaccine-preventable, acute, and chronic. Columns represent PPH rates of each diseases category for males, females, and total for both males and females. PPH, potentially preventable hospitalisation; *JKN*, *Jaminan Kesehatan Nasional* (Indonesia's national health insurance scheme).

### Variation in PPH across districts/cities

We observed wide variation in age-standardised overall PPH rates across 513 districts/cities, grouped into seven regions (**Figure 2, Supplemental table 2**). Rates ranged from as low as 0.3 per 10,000 in Memberamo Tengah District (Papua) to 1,054.8 per 10,000 in Lhokseumawe City (Sumatera). Relatively high rates (above 100 per 10,000) were seen in 88% of districts/cities in Java-Bali, 80% in Sulawesi, 78% in Sumatera, and 70% in Kalimantan, while no districts/cities in these regions recorded rates below 15 per 10,000. In contrast, the rates were generally lower in districts/cities in Papua, where over one-third of districts/cities recorded rates lower than 15 per 10,000, including some districts which had rates below 5 per 10,000 (22%) or close to zero (5%). Nusa Tenggara and Maluku showed intermediate patterns with around half of districts/cities in these regions, with rates above 100 per 10,000, while the lowest rates observed in these regions (14.8 and 7.2 per 10,000, respectively) were still higher than those observed in Papua.

**Figure 2.**
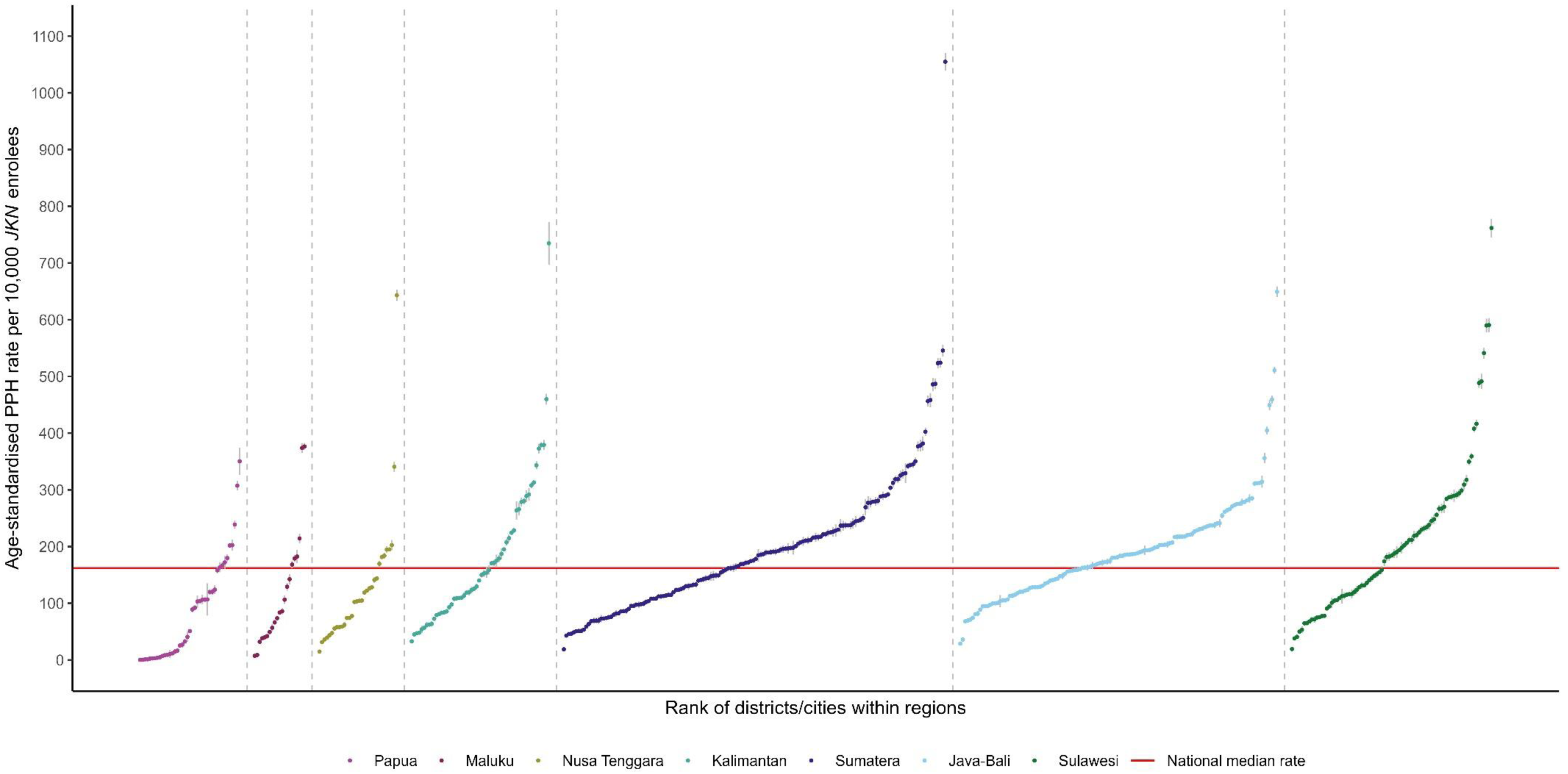
Distribution of age-standardised overall PPH rates per 10,000 *JKN* enrolees across districts/cities, grouped by region Each point represents one district/city, with error bars indicating 95% confidence intervals. Overall PPH rates were age-standardised, calculated using direct age standardisation with national *JKN* enrolees as the standard population. Sampling weights provided in NHISD 2023 were applied. Our analysis included 513 of 514 districts/cities, excluding *Deiyai* District in Papua due to the absence of *JKN*-contracted hospitals in 2023. Districts/cities were grouped into seven regions for interpretability, were ranked from lowest to highest rates within the region and were colour-coded by region. Regions were ordered from left to right by the lowest median age-standardised overall PPH rates across seven regions (Papua=51.0, Maluku=83.7, Nusa Tenggara=103.9, Kalimantan=145.1, Sumatera=176.8, Java-Bali= 182.4, and Sulawesi=184.4 per 10,000 *JKN* enrolees). The red horizontal line indicates the national median age-standardised overall PPH rate (162.0 per 10,000 *JKN* enrolees). PPH, potentially preventable hospitalisations; *JKN*, *Jaminan Kesehatan Nasional* (Indonesia’s Social Health Insurance); NHISD, National Health Insurance Sample Data.

In the multilevel models, results from model 1 confirmed variation in PPH (overall and by type) across districts/cities (**Table 3**). The MRRs for overall, vaccine-preventable, acute and chronic PPH were 1.62, 1.83, 1.72, 1.53, respectively. This indicates that individuals living in a district/city with a higher-than-average PPH rate had a 53−83% greater likelihood of experiencing above-average PPH compared to individuals with characteristics living in lower-rate areas. Adding sociodemographic characteristics (age, marital status, sex, and type of *JKN* membership) in model 2 showed that variation in PPH still remained with MRRs of 1.52, 1.82, 1.61, and 1.41, respectively and explained only a small part of the geographic variation in PPH.

**Table 3.** Multilevel model results: MRRs and percentage of reduction in MRRs between models.

| Outcomes (PPH) | Model 1 | Model 2 | MRR reduction (%)<br>Model 1 to 2 |
| --- | --- | --- | --- |
| Overall | 1.62 | 1.52 | 6.25 |
| Vaccine-preventable | 1.83 | 1.82 | 0.59 |
| Acute | 1.72 | 1.61 | 6.53 |
| Chronic | 1.53 | 1.41 | 7.44 |

Models were estimated from multilevel negative binomial regression.
Model 1: null model with random intercept for district/city.
Model 2: adjusted for individual sociodemographic characteristics (age, marital status, sex, and type of *JKN* membership).
Percentage of reduction in MRRs from model 1 to model 2 = $(\text{MRR}_{\text{Model 1}} - \text{MRR}_{\text{Model 2}} / \text{MRR}_{\text{Model 1}}) \times 100$
PPH, potentially preventable hospitalisation; MRR, median rate ratio.

### Association between sociodemographic characteristics and PPH

PPHs were associated with age, marital status, sex, and type of *JKN* membership (**Figure 3, Supplemental table 3**). Compared to young adults (25−34 years), older adults were more likely to have PPH, particularly chronic (55−64 years IRR=12.05, 95% CI 11.25−12.91 and 65−74 years IRR=13.66, 95% CI 12.69−14.70), while young age (0−14 years) were more likely to have vaccine-preventable and acute PPH (IRR=5.22, 95% CI 4.02−6.79 and IRR=5.45, 95% CI 5.18−5.73, respectively). Being married or divorced was modestly associated with higher acute, chronic, and overall PPH than being single. We observed similar pattern of associations for type of *JKN* membership with all PPH types for non-subsidised *JKN* enrolees exceeding those for subsidised group. Females were slightly more likely than males to have acute and overall PPH, and less likely to have vaccine-preventable and chronic PPH.

**Figure 3.**
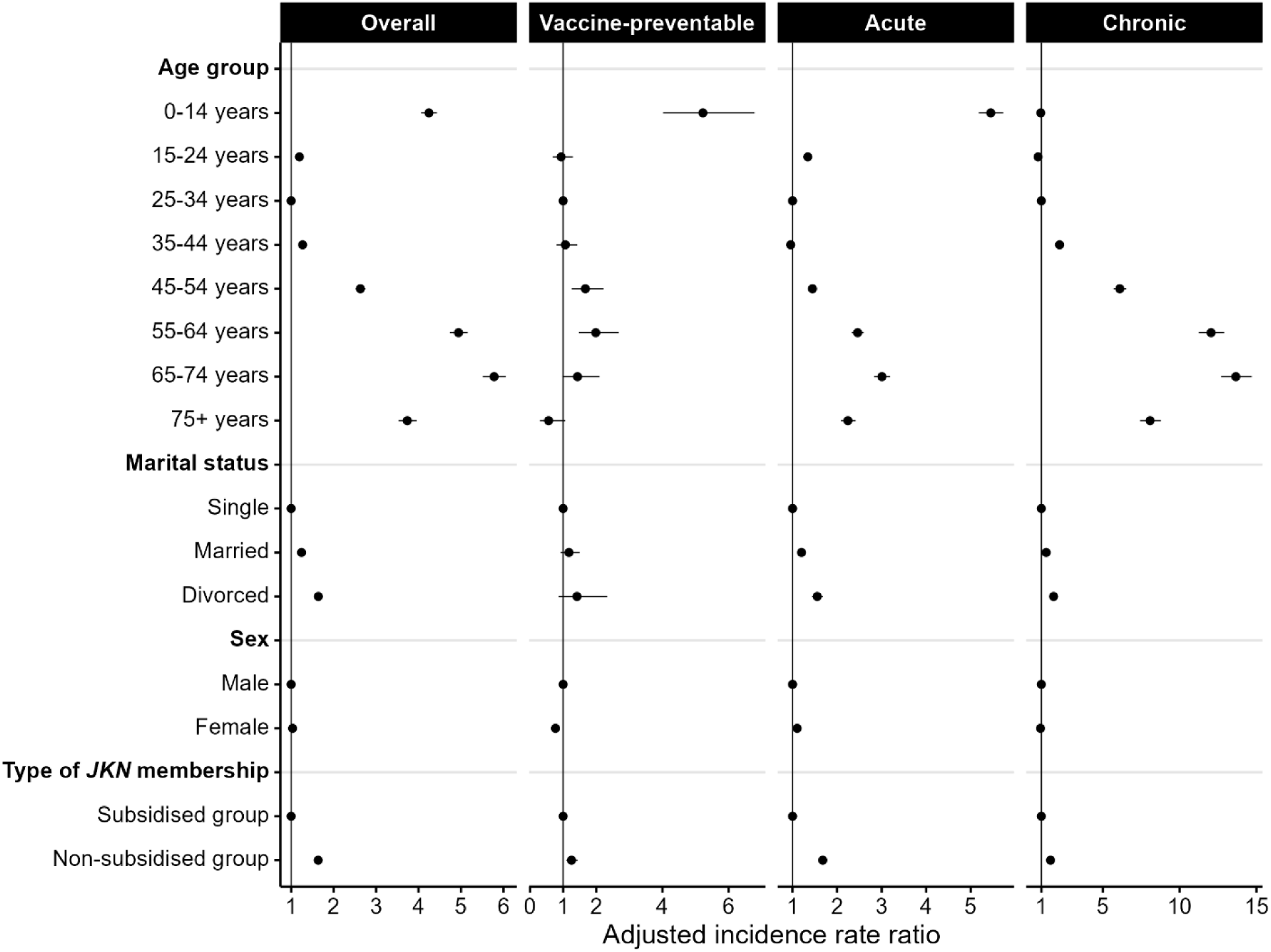
Multilevel model results: IRRs (95% CIs) for associations between sociodemographic characteristics and PPHs (overall and by type: vaccine-preventable, acute, and chronic) IRRs (95% CIs) for PPHs were estimated using multilevel negative binomial regression, mutually adjusted for individual sociodemographic characteristics (age group, marital status, sex, and type of *JKN* membership). IRR, incidence rate ratio; CI, confidence interval; PPH, potentially preventable hospitalisations.

## DISCUSSION

### Summary of the principal findings

To our knowledge, this study provides the first national assessment of PPH among *JKN* enrolees in Indonesia, highlighting variations across districts/cities. In 2023, the overall PPH rate was 180.9 per 10,000 *JKN* enrolees, with rates slightly above 90 per 10,000 for both acute and chronic PPH, and less than 3 per 10,000 for vaccine-preventable PPH. Here, most districts/cities in Java-Bali, Sulawesi, Sumatera, and Kalimantan reported highest overall PPH rates. Nusa Tenggara and Maluku had lower rates, while Papua recorded the lowest rates. Notably, these areas with low PPH rates experience greater socioeconomic disadvantage and have more limited availability and accessibility of hospitals services than other areas. Variation in PPH across districts/cities was evident even after adjusting for sociodemographic characteristics. Strong associations with higher PPH were seen for older adults 55−64 and 65−74 years (particularly chronic) and young age 0−14 years (vaccine-preventable and acute) compared to young adults (25−34 years). Modest associations with higher acute, chronic, and overall PPH were observed for those married or divorced compared with being single, and across all PPH types for those with non-subsidised *JKN* enrolees, whereas sex made little difference.

### Comparison with existing literature

The leading PPH conditions in Indonesia (dehydration and gastroenteritis, bacterial pneumonia, diabetes and associated complications, hypertensive complications, cerebrovascular disease, and tuberculosis) broadly align with those reported in a study from Vietnam [23]. However, a notable difference is that Vietnam reported higher PPH rates for ear, nose, throat (ENT) and chronic obstructive pulmonary disease (COPD), whereas tuberculosis was a more prominent contributor to PPH in Indonesia.

Geographic variation in PPH has been well-documented internationally [31–35] and we observed this variation in Indonesia. At the area-level, however, we found that lower PPH rates among most districts/cities in Papua, where almost half of Indonesia’s disadvantaged districts/cities are located [36], was unexpected. Health outcomes in most districts/cities in this region remain among the poorest nationally, including one of the lowest life expectancies [37], the highest prevalence of malaria [26], high burdens of diarrhoea and pneumonia [38], and tuberculosis [39] and they were expected to experience higher PPH. These findings seemingly contradict evidence from high-income countries, such as Switzerland [33] and Australia [35], where higher PPH rates are typically reported in more deprived areas. Possibly, this can be explained by the fact that in Indonesia where hospital access is far from universal, particularly in disadvantaged districts/cities like those in Papua, low PPH rates may indicate limited hospital availability and accessibility, rather than better PHC quality. This interpretation is supported by previous evidence showing that hospital inpatient utilisations in Papua were the lowest nationally [40], likely due to low hospital bed ratio [41], limited health workforces availability [42] and geographic barriers to access hospital care, particularly longer travel time and high transportation cost [43].

Our findings highlight the important sociodemographic differences in PPH, particularly regarding type of *JKN* membership. Non-subsidised *JKN* enrolees had higher PPHs, across all types, than subsidised members who are socioeconomically disadvantaged. This also contrasts with findings from high-income countries, where higher PPH are more common among lower socioeconomic groups [32,44]. In Indonesia, this may be explained by greater access to hospital services among non-subsidised *JKN* enrolees, including the more ability to cover transportation cost, more aware of health issues, and more informed regarding the health insurance benefits, thus more likely to utilise inpatient hospital services and to experience higher PPH than subsidised *JKN* enrolees (lower socioeconomic group). This interpretation is consistent with a recent study on hospital utilisation in Indonesia which found that non-subsidised enrolees were more likely to utilise hospitals than subsidised and uninsured groups [45].

Our other sociodemographic characteristics findings indicate that children were more likely to experience vaccine-preventable and acute PPH which is comparable to existing international studies [46,47]. These findings reflect Indonesia’s ongoing challenges in achieving universal immunisation coverage [48] and in addressing acute conditions for children, particularly acute respiratory infection and diarrhoea which remain common among children living with poor sanitation and malnutrition [49]. Older people were more likely to have chronic PPH, a pattern that has been previously observed internationally [50–53]. This is consistent with the existing evidence that chronic conditions are the leading contributors to inpatient care utilisations in older Indonesians [54,55]. Additionally, our findings that females were slightly higher acute and overall PPH than males and those not single were more likely to have acute, chronic, and overall PPH, may be reflect their greater hospital utilisations, including inpatient services, as reported in a previous study conducted in Indonesia [28].

Our findings showing that individual sociodemographic characteristics partly explained geographic variation in PPH are in line with previous studies in high-income countries [32,53]. However, our study included only a limited set of sociodemographic variables, and many important socioeconomic characteristics, health factors, and behaviours were not available in the NHISD dataset. It, therefore, remains uncertain to what extent geographic variation could be further explained if more complete data were available.

### Implication to policy, practice, and future research

Our findings have important implications for the Government of Indonesia’s ongoing efforts to achieve equitable access to high-quality PHC. Many of the most common conditions contributing to PPH in Indonesia were acute and chronic conditions that are largely preventable through improved hygiene (e.g. dehydration and gastroenteritis, bacterial pneumonia) or through health promotion, early case detection, prompt and effective treatment (e.g. diabetes and associated complications, hypertensive conditions, cerebrovascular disease, tuberculosis). These findings may reflect weaknesses in PHC in Indonesia, particularly in preventive and promotive programmes as well as effective treatments. Strengthening PHC capacity to prevent, detect, and manage the conditions identified as leading contributors to PPH in this study across districts/cities may not only reduce geographic variation in PPH, but also improve the efficiency of *JKN* resource use. This could also help *BPJS Kesehatan* mitigate potential future financial shortfalls and maintain sustainability of *JKN* resources.

Although PPH has potential as a measure of PHC quality in Indonesia, policymakers should interpret it cautiously. In disadvantaged districts/cities, where hospital availability and accessibility remain limited, lower PPH may reflect barriers to access hospital care rather than better PHC quality. Without addressing these gaps, PPH may not properly capture PHC quality. To support routine monitoring of PHC quality, establishing an Indonesia-specific list of conditions to define PPH would help ensure that the measure fully reflects the local context. The list used in this study could serve as a starting point, which the Government of Indonesia might refine through consultations with clinicians, public health experts, and policymakers to assess the relevance of each condition and its applicability for national PHC quality measurement.

A substantial proportion of geographic variation in PPH remained unexplained, likely underscoring the need for greater understanding of unmeasured factors in our study at both the individual-level (e.g. socioeconomic characteristics such as education, employment; health factors such as self-rated health, adherence to medications, severity of illness; and health behaviours such as health care seeking behaviour, smoking, physical activity) and area-level (e.g. PHC and hospitals service characteristics) that potentially affecting geographic variation in PPH [56]. Future studies need to explore these factors to better clarify the drivers of geographic variation in PPH in Indonesia. In addition, strengthening data systems to enable linkage between administrative datasets (NHISD) and other health or socioeconomic survey datasets at the individual-level would provide a more comprehensive assessment of PPH in the country. Developing such capacity in Indonesia would strengthen the evidence base to inform responsive PHC policies, which is currently not feasible.

### Strengths and limitations of this study

This study represents the first national assessment of PPH in Indonesia and provides new perspectives on PHC quality measurement using PPH, which has been extensively studied in high-income settings. We used a large-scale, nationally representative, and routinely collected sample dataset from the *JKN* program, which is both up-to-date and one of the most comprehensive sources of anonymised individual-level healthcare utilisation records available. As we applied sampling weights, findings from this study can be generalisable to Indonesia’s *JKN* enrolees, which reached almost the entire Indonesian population (95.2% in 2023 [57]). We also employed multilevel analysis, which allowed us to account for the hierarchical structure of the data, individuals nested within districts/cities.

However, some limitations should be noted. Not all healthcare providers were contracted to provide health services under the *JKN* scheme in 2023. To participate, providers must meet accreditation and other requirements. Some private providers, however, have operated independently outside the *JKN* scheme. According to a report from national statistics bureau, approximately 28% of hospitalisations in 2023 were not using *JKN* nationally [58]. These hospitalisations were therefore not captured in the data that we used for this study, although a fair proportion of them likely involved elective care rather than conditions classified as PPH. Thus, our study still captured the majority of PPH nationally. Additionally, to analyse PPH in Indonesia, we modified list of conditions from other countries, which may not fully reflect the Indonesian context, as the country has not yet developed its own official list of conditions for defining PPH.

## CONCLUSION

This study highlights that PPH varied across districts/cities in Indonesia, even after adjusting for a range of sociodemographic characteristics. Given Indonesia’s uneven access to hospital services, caution is required when interpreting PPH as a measure of PHC quality, particularly in disadvantaged areas, such as districts/cities in Papua. Lower estimates may reflect a lack of availability and accessibility to hospital services rather than better PHC quality. This underscores the need for context-specific interpretation when using PPH to assess PHC quality.

## Data Availability

This study used official government data which are publicly available through request to *BPJS Kesehatan* (https://data.bpjs-kesehatan.go.id/bpjs-portal/action/datasample.cbi).

## Acknowledgement

The authors would like to thank *BPJS Kesehatan* for providing anonymised secondary datasets used in this study.

## Author Contributions

DAS conceived and designed this study, completed analysis, drafted and finalised the manuscript, and acted as guarantor. DCB, RJK, and MK supervised and reviewed the study design, analysis, and manuscript. All authors revised the work for intellectual content and approved the final version of the manuscript.

## Patient and public involvement

Patients and/or the public were not involved in the design, or conduct, or reporting, or dissemination plans of this study.

## Patient consent for publication

Not applicable.

## Ethics approval

Ethics approval for this study was obtained from The Australian National University Human Research Ethics Committee (H/2024/1199).

